# Measuring wealth in rural and urban Africa: findings and recommendations from a multi-country study

**DOI:** 10.1101/2025.04.30.25326739

**Authors:** Anthony Muchai Manyara, Momodou Jallow, Tadios Manyanga, Etheldreda I Yoliswa Madela, Fundile Habana, Nyasha Buwu, Awa Touray, Chris Grundy, Hannah Wilson, Anya Burton, Bilkish Cassim, Rashida A Ferrand, Kate A Ward, Celia L Gregson

## Abstract

Wealth indices enable socioeconomic status measurement, particularly in low- and middle-income countries. However, their reliability, validity, and data collection and computation efficiency can be improved. This analysis aimed to generate four wealth indices in five geographical settings (rural and urban Gambia, rural and urban South Africa, and urban Zimbabwe), to compare and validate these indices and formulate recommendations for wealth measurement in Africa. Population-based cross-sectional data from 5,296 adults aged ≥40 years were analysed. Principal Component Analyses generated four wealth indices based on 18 variables quantifying house ownership, wall and roofing material, and asset ownership: (1) geographical site-specific, (2) national-level, (3) national composite, and (4) ‘international’ wealth indices. Recommendations were based on reflective workshops (totalling 6 hours) held with experienced in-country fieldwork coordinators. A very strong correlation (>0.9) between the national-level, national composite and ‘international’ indices was seen, with strong correlations (>0.6) between these three indices and the site-specific index. In all sites, the ‘international’ wealth index was associated with greater asset ownership in 44.4% (8/18) of the variables intended to measure wealth *i.e*., refrigerator, television, working car/truck, tap in the house, flush toilet, decoder/satellite dish, computer, and tiled floors, albeit with rural-urban and country differences. The ‘international’ wealth index had a moderate prediction (>0.55) across all the other SES measures: higher household income, greater educational attainment, and food security. Reflective workshops established that the accuracy of wealth measurement for multi-country and multi-site studies, using asset-based indices, needs an unambiguous, differential, and clearly defined asset list. Furthermore, consulting local teams to select these assets can reduce data collection burden while increasing index validity, including ‘contemporary’ asset assessment, e.g., Wi-Fi internet connection. Finally, it is recommended that generated wealth indices be routinely internally validated against individual asset ownership and other socioeconomic measures such as educational attainment.

## Introduction

Socioeconomic status (SES) is a major social determinant of health(1, 2), and SES is frequently considered a confounder in exposure-outcome associations(3). Consequently, SES data are widely collected in health research(3). Income and expenditure can be used as SES measures, but their data collection can be time and resource-intensive, and in low-income settings reliability and validity are challenged by informal, diverse, and variable ‘unofficial’ income sources(3–5). Wealth indices, based on ownership of household assets or other relevant data (e.g., housing construction material, water and sanitation facilities), provide a more valid measure of SES in such settings, potentially reflecting both income and expenditure(3, 6). Use of wealth indices in low-income countries was pioneered by Demographic Health Surveys (DHS)(4, 7), nationally representative surveys conducted in more than 85 countries for over three decades, mainly focusing on maternal and child health(8). The DHS wealth index and other indices are composite scores based on ownership of household assets (e.g., having a television), materials used to construct houses, and access to water and sanitation facilities(7). While such indices have been extensively used (as predictors or confounders) in studies that measure health or social outcomes, research into their reliability, validity, and optimisation is limited but emerging(6).

One of the limitations of the DHS wealth index is its ‘urban bias’: assigning more weight to assets owned in urban settings, e.g., television, electricity(9, 10). Martel and co-authors recently proposed a new approach (polychoric dual-component wealth index) to reduce ‘urban bias’(9). Secondly, while data needed to generate such indices may be ‘easy’ to collect (simple yes/no responses), given the 30-100 variables needed for index generation, it conveys a substantial data collection and computation burden(11, 12). Xie et al recently proposed a method (feature selection principal component analysis) to create a wealth index for Mozambique that reduces this burden(10). Thirdly, the reliability of wealth indices has been challenged after Smith et al found that interviewing different household members (husbands versus wives) produced different asset ownership scores, in rural Uganda(5). Fourthly, using different assets to generate wealth indices can limit comparability between sites and countries. Consequently, the International Wealth Index (IWI) was developed to improve comparability(13)

We proposed to investigate the reliability and validity of wealth indices in Africa across three countries based on a study of rural and urban settings in Gambia and South Africa and an urban setting in Zimbabwe. Specifically, this analysis aimed to firstly describe the generation of wealth indices, including validation against household income, food insecurity and educational attainment, and secondly make practical recommendations for wealth index data collection and analysis, based on consultations with experienced in-country fieldwork coordination teams from each country.

## Methods

### Study design, population and recruitment

This study used data from a population-based cross-sectional survey whose primary aim was to determine the prevalence of vertebral fractures and musculoskeletal multimorbidity as part of the Fractures-E3 (Fractures in sub-Saharan Africa: epidemiology, economic impact, and ethnography) research programme, whose protocol has previously been published(14). The survey aimed to recruit men and women aged ≥40 years across five geographical sites: rural and urban Gambia, rural and urban South Africa, and urban Zimbabwe between 2022 and 2024. The target sample size was 504 women and 504 men in each of the five geographical sites (total=5,040), evenly distributed (n=168) across 3 age-strata (40-54, 55-69 and ≥70 years) providing power to detect an odds ratio of 2.0 for risks with ≥13% prevalence and outcomes with ≥9% prevalence(14).

Each of the five geographical sites was divided into smaller blocks using satellite imagery and OpenStreetMap within the QGIS Geographic Information System software. These blocks were designed to be approximately equal in size and sufficiently large to recruit 18 to 30 eligible participants, i.e., 3 to 5 individuals per age-sex category. Each block was assigned a random number, allowing for the random selection of a subset. In each selected block, all households were visited, enumerated, and eligible individuals were invited to participate until the required age-sex strata were filled.

### Data collection and definitions

Standardised and validated procedures were used to collect data: researcher-administered questionnaires, and clinical and physical assessments as detailed in the protocol(14). Relevant to this analysis, a researcher-administered questionnaire collecting 18 variables (see Wealth Indices Generation) describing asset ownership, housing, wall and roofing materials, and these variables were used to compute the wealth index. Other SES measures were collected: household income, participant-level educational attainment, and food insecurity. Food insecurity was determined using five *Yes/No* questions taken from the Household Food Insecurity Access Scale, capturing food insufficiency in the last four weeks(15). Household income was self-reported using pre-determined categories and defined for analyses as no income; low-income (≤4000 Gambian Dalasi, ≤100 US dollars in Zimbabwe, or ≤1000 South African Rand); middle-income (40,001-16,000 Gambian Dalasi, 101-500 US dollars in Zimbabwe, 1,001-3,000 South African Rand); and high-income (>16,000 Gambian Dalasi, >500 US dollars in Zimbabwe, >3,000 South African Rand). Educational attainment was categorised as having no formal education, primary, or secondary and above. Food insecurity was categorised as food secure (*No* to all five questions), moderately food insecure (*Yes* to 1-2 questions), or severely food insecure (*Yes* to 3-5 questions).

### Wealth indices generation

Principal components analyses (PCA) were used to generate four wealth indices: (1) site-specific, (2) national, and (3) ‘international’ indices, with a further index (4) national composite, computed using site-specific and national-level indices, as described below. Table 1 shows the 18 variables used to generate each of the indices. All analyses were performed using R (version 4.2.0) software, with PCA using the ‘psych’ package(16).

**Table 1:**
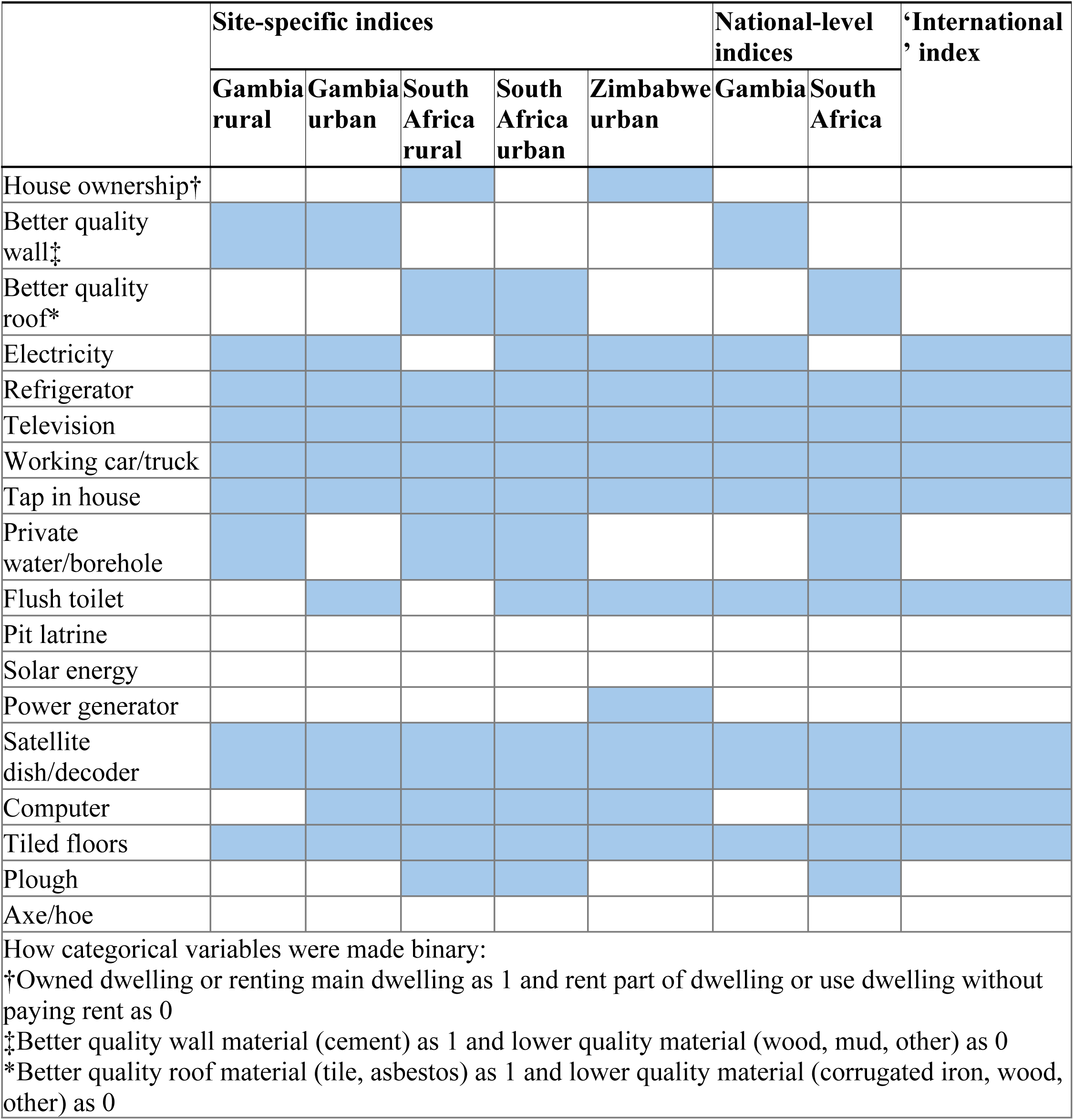
Assets used (in dark shading) in final PCA for the different indices.

### The site-specific wealth index

The PCA was an iterative process with the first analysis using all 18 variables and then sequentially eliminating variables. A second analysis was based on fewer variables after elimination based on theoretical considerations such as the Kaiser-Meyer-Olkin index to check for sampling adequacy(17). In-country teams were then consulted on these eliminated variables to verify the appropriateness of their elimination, according to their opinion on the validity of the variable as a measure of wealth; and then similarly verified the appropriateness of retaining the other asset variables. If further elimination was needed, a further analysis was performed to improve PCA model characteristics (Table 2).

**Table 2:**
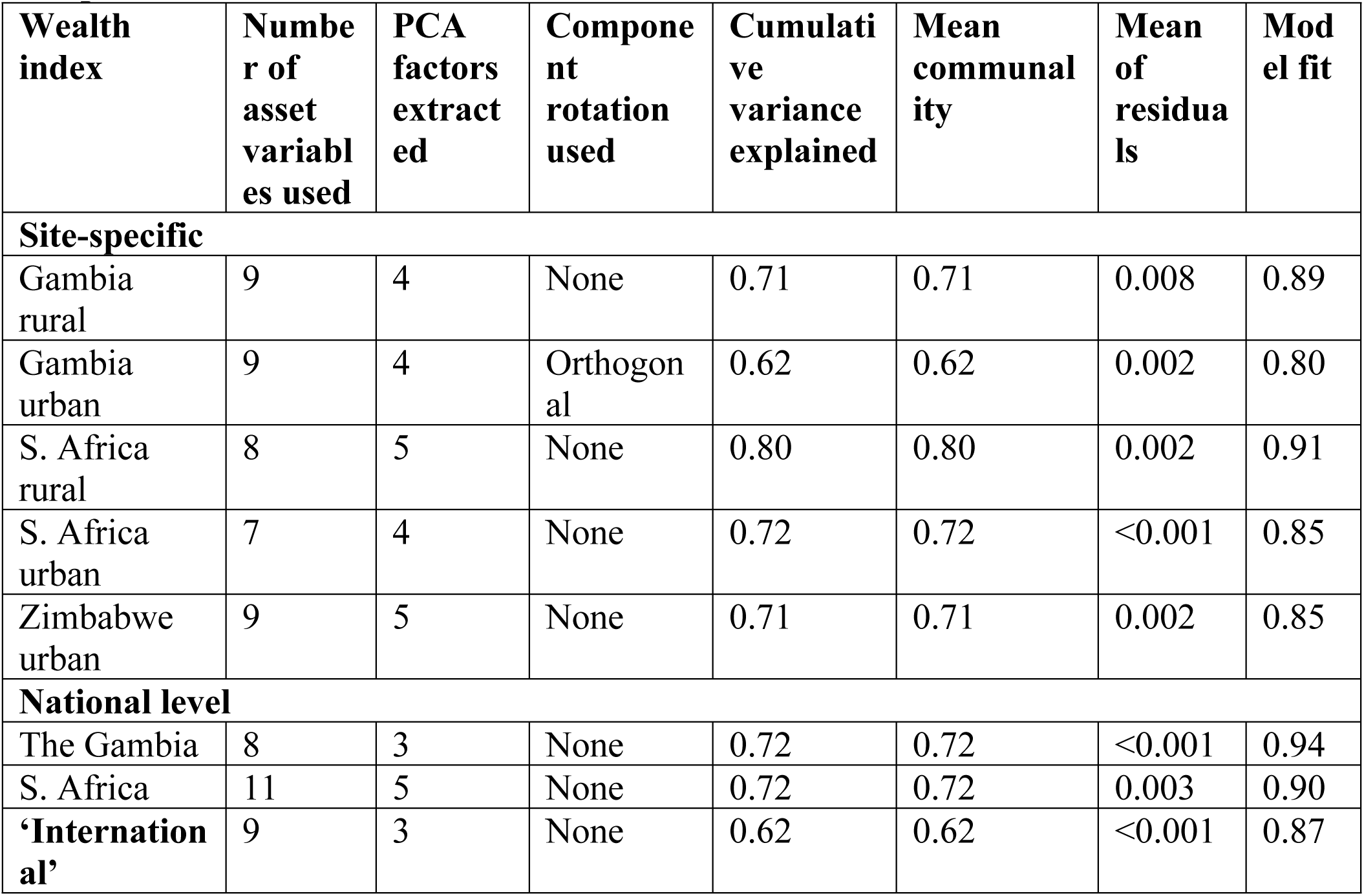
Characteristics of the principal component analysis (PCA) used to generate the site-specific, national-level, and international wealth indices.

The lead author (AMM) facilitated two one-hour reflective workshops (six in total) with in-country field team leads, which involved MJ and AT from The Gambia, TM and NB from Zimbabwe, and YM and FH from South Africa. The workshops were held virtually with notes taken (by the facilitator) to capture structured discussions. Issues discussed included: limitations of variables used in the analyses; face validity of trends observed with wealth index tertiles; and other assets that can be used to measure wealth The final and refined site-specific wealth index was then categorised into tertiles corresponding to low, middle, and high wealth. To check the index’s face validity, the distribution of the 18 asset variables was examined across the tertiles.

### National, composite, and ‘international’ indices

To enable comparability of the wealth indices across the sites and countries, national-level, national composite and ‘international’ indices were computed using DHS approaches(18). First, a national-level wealth index for The Gambia and South Africa was generated using variables that were good measures of wealth in both rural and urban sites i.e., ownership of the asset increased with site-specific wealth tertiles, see Table 1. Second, national composite indices for The Gambia and South Africa were computed by regressing urban and rural indices separately against the national-level index. Then using the formula y=mx+c rural and urban indices were computed, which were combined to a national composite index(18).

Textbox 1 shows the coefficients used in the formula for the four sites.

#### Textbox 1: Formulae used to generate national composite wealth indices for the Gambia and South Africa

**Gambian composite wealth index**

Rural: -0.372981 + 0.630133 * rural-specific index

Urban: 0.323228 + 0.565765 * urban-specific index

**South African composite wealth index**

Rural: -0.296153 + 0.780544 * rural-specific index

Urban: 0.290972 + 0.796054 * urban-specific index

Finally, an ’international’ wealth index was generated using variables that were good measures of wealth based on The Gambian and South African national-level indices and the Zimbabwean urban index, see Table 1.

### Characteristics of PCA models used

The number of assets used in the PCAs ranged from 7-12 with the number of principal components extracted ranging from 3-5 (Table 2). Orthogonal and oblique rotations(17) were attempted in each PCA but were dropped if the resultant wealth index was skewed or lacked face validity. The cumulative variance explained by the extracted principal components was high, ranging between 62-80%, with the mean of the residuals being acceptable as they were well below the recommended threshold of <0.08(17, 19). The mean communality (shared variance among the variables) was between 0.62-0.80 which is within the acceptable cutoff of ≥0.6(17). Model fit was good, ranging between 80-95% (Table 2).

### Comparison of indices

Comparisons between the four indices were performed using a correlation matrix. Smoothed kernel density plots were used to visualise the four indices across the geographical sites. Mean ± standard deviation, median (interquartile range), minimum and maximum values, and percentiles of the indices in all sites were computed. The predictive performance of each of the four indices was evaluated against self-reported household income, educational attainment, and food insecurity. Logistic regression with 10-fold cross-validation (nine data parts used for training and one part for testing) was used with the indices as continuous variables (predictor) and household income, educational attainment, and food insecurity as binary variables (outcomes). High household income was defined as middle and high-income categories (described above), high educational attainment was defined as having secondary or tertiary education, and food security was defined as *No* to all five questions described above.

### Internal validation

The final wealth index was categorised into tertiles separately in each site, to provide sufficient numbers in each tertile. In rural Gambia, ranked perturbed probabilities (probabilities are slightly adjusted (perturbed) and then ranked in order) were used to generate tertiles given duplicate breakpoints. To internally validate this wealth index, the distributions of the 18 asset variables were compared across the wealth tertiles. Furthermore, the distributions of self-reported household income, educational attainment, and food insecurity were compared across wealth tertiles. Pearson’s Chi-square tests were used to assess associations between wealth index tertiles and ownership of assets, household income, educational attainment and food insecurity categories.

### Reflective workshops

As described above, reflective workshops contributed to refinement and face validation of the site-specific wealth index. Moreover, the workshops explored the strengths and limitations of variables used to measure wealth in the current study including household income. Proposals were made for other variables that could be potential measures of wealth for future studies. Simple thematic analysis was used to draw out themes from these discussions, which were synthesised as text.

## Results

### Sociodemographic characteristics

In total, 12185 households were visited, identifying 8840 eligible individuals, of whom 6481 were invited to participate. Of those invited, 5296 (81.7%) agreed, attended the local research clinic, and provided informed consent or assent (either directly or via proxy) to participate. These participants are included in the analysis (see S1 Appendix for participant flow diagram).

Across the three countries, 2270 (83.2%) were recruited from The Gambia, 1916(89.8%) from South Africa and 1110 (68.6%) from Zimbabwe, S1 Appendix. The total study population of 5296 had a mean (SD) age of 61.1 (12.9) years, and 51.6-56.1% were female (Table 3). Being single (including separated and divorced) was more common in South Africa and rare in The Gambia, while low educational attainment was more common in The Gambia and rare in South Africa and Zimbabwe. Self-report of ‘No income’ was highest in Zimbabwe. Food insecurity was more common in South Africa than in The Gambia and Zimbabwe (Table 3).

**Table 3:** Sociodemographic characteristics of participants across the five study sites.

| **Characteristic** | **Overall, n=5296** | **Gambia Urban, n=1218** | **Gambia rural, n=1052** | **SA Urban, n=968** | **SA rural, n=948** | **Zimbabwe, n=1110** |
| --- | --- | --- | --- | --- | --- | --- |
| **Age – Mean±SD** | 61.0±12.9 | 60.6±12.9 | 61.5±13.2 | 60.1±11.9 | 60.2±11.9 | 62.5±14.1 |
| **Sex** |  |  |  |  |  |  |
| Female | 2,823 (53.3%) | 671 (55.1%) | 547 (52.0%) | 520 (53.7%) | 513 (54.1%) | 572 (51.5%) |
| Male | 2,473 (46.7%) | 547 (44.9%) | 505 (48.0%) | 448 (46.3%) | 435 (45.9%) | 538 (48.5%) |
| **Marital status** |  |  |  |  |  |  |
| Single | 1,248 (23.6%) | 55 (4.5%) | 30 (2.9%) | 559 (58.0%) | 442 (46.7%) | 162 (14.6%) |
| Married | 2,976 (56.3%) | 923 (76.0%) | 868 (82.5%) | 260 (27.0%) | 345 (36.4%) | 580 (52.3%) |
| Widowed | 1,062 (20.1%) | 236 (19.4%) | 154 (14.6%) | 144 (15.0%) | 160 (16.9%) | 368 (33.2%) |
| **Household income** |  |  |  |  |  |  |
| No income | 360 (7.3%) | 50 (5.3%) | 0 (0.0%) | 10 (1.1%) | 18 (1.9%) | 282 (25.6%) |
| Low-income | 1,166 (23.6%) | 325 (34.3%) | 246 (23.5%) | 82 (9.1%) | 30 (3.2%) | 483 (43.9%) |
| Middle-income | 2,506 (50.8%) | 508 (53.6%) | 749 (71.6%) | 494 (54.9%) | 442 (47.0%) | 313 (28.5%) |

| Characteristic | Overall,<br>n=5296 | Gambia<br>Urban, n=1218 | Gambia<br>rural, n=1052 | SA Urban,<br>n=968 | SA rural,<br>n=948 | Zimbabwe,<br>n=1110 |
| --- | --- | --- | --- | --- | --- | --- |
| High-income | 900 (18.2%) | 64 (6.8%) | 51 (4.9%) | 313 (34.8%) | 450 (47.9%) | 22 (2.0%) |
| **Educational attainment** |  |  |  |  |  |  |
| No Education | 1,777 (33.9%) | 776 (65.3%) | 836 (80.0%) | 21 (2.2%) | 59 (6.2%) | 85 (7.7%) |
| Primary | 1,207 (23.0%) | 137 (11.5%) | 95 (9.1%) | 218 (22.6%) | 445 (47.0%) | 312 (28.3%) |
| Secondary and above | 2,262 (43.1%) | 276 (23.2%) | 114 (10.9%) | 724 (75.2%) | 442 (46.7%) | 706 (64.0%) |
| **Food insecurity** |  |  |  |  |  |  |
| Food secure | 2,939 (55.6%) | 826 (68.0%) | 666 (63.3%) | 308 (31.9%) | 374 (39.5%) | 765 (68.9%) |
| Moderate food insecure | 682 (12.9%) | 202 (16.6%) | 165 (15.7%) | 124 (12.8%) | 129 (13.6%) | 62 (5.6%) |
| Severely food insecure | 1,668 (31.5%) | 187 (15.4%) | 221 (21.0%) | 533 (55.2%) | 444 (46.9%) | 283 (25.5%) |
| SD – Standard deviation |  |  |  |  |  |  |
| †Low-income ($\leq 4000$ Gambian Dalasi, $\leq 100$ US dollars in Zimbabwe, $\leq 1000$ South African Rand); middle-income (40,001-16,000 Gambian Dalasi, \$101-500 US dollars in Zimbabwe, R1,001-3,000 South African Rand); and high-income ($> 16,000$ Gambian Dalasi, $> \$500$ US dollars in Zimbabwe, $> R3,000$ South African Rand) | | | | | | |

### Comparison and validation of wealth indices

The site-specific and ‘international’ wealth indices in urban Zimbabwe were normally distributed (Figure 1).

**Figure 1:**
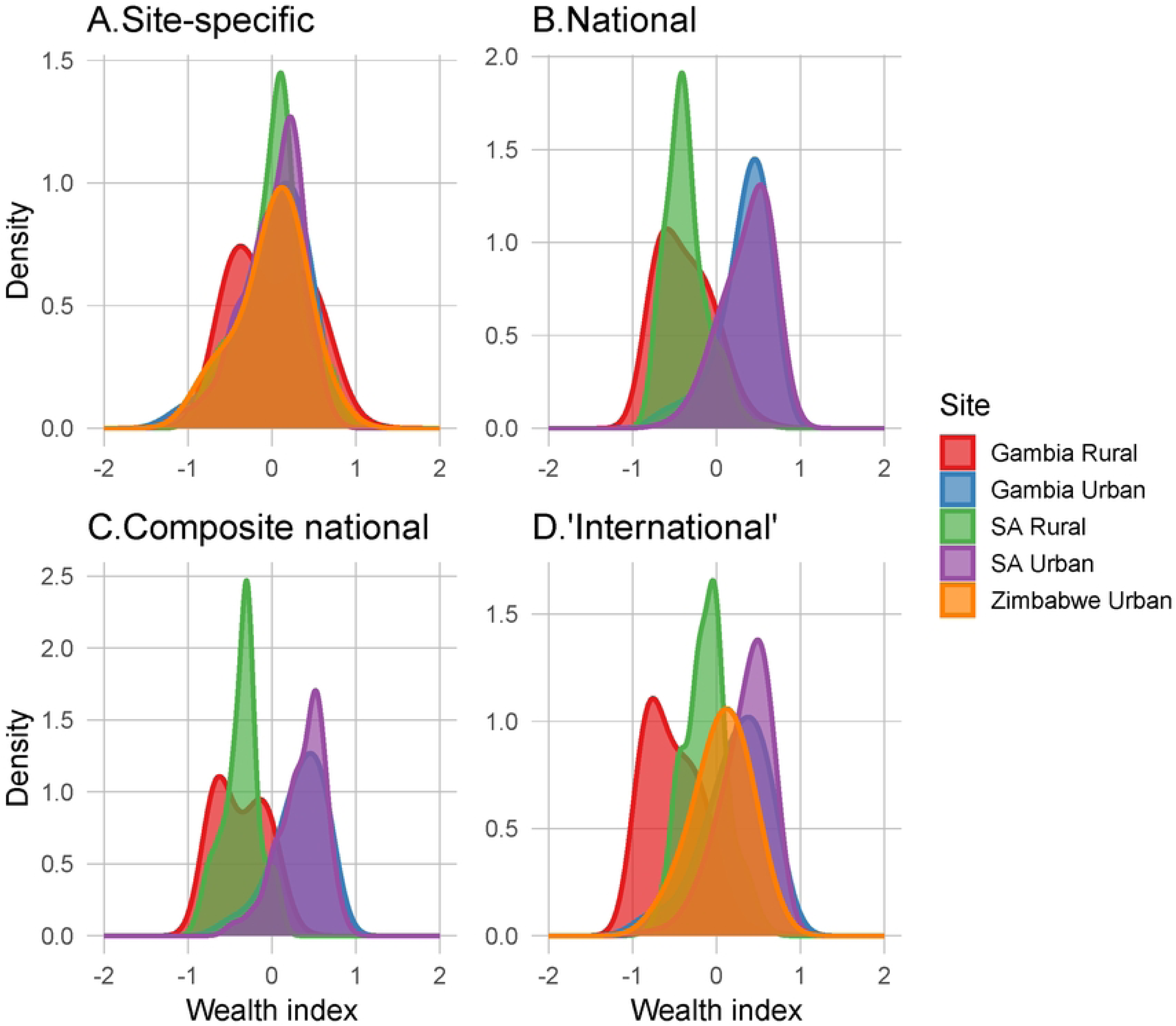
Smoothed kernel density plots of the four wealth indices across the five geographical study sites

Zimbabwe lacked national-level and composite indices, as data were only collected from one (urban) site. In The Gambia and South Africa, all wealth indices were skewed, with urban sites to the right (i.e., more wealth) and rural sites to the left (i.e., less wealth), with urban South Africa and rural Gambia being to the extreme ends, respectively.

Very strong correlations (>0.9) were seen between the national-level, national composite and ‘international’ indices, with strong correlations (>0.6) between these indices and the site-specific index (Figure 2).

**Figure 2:**
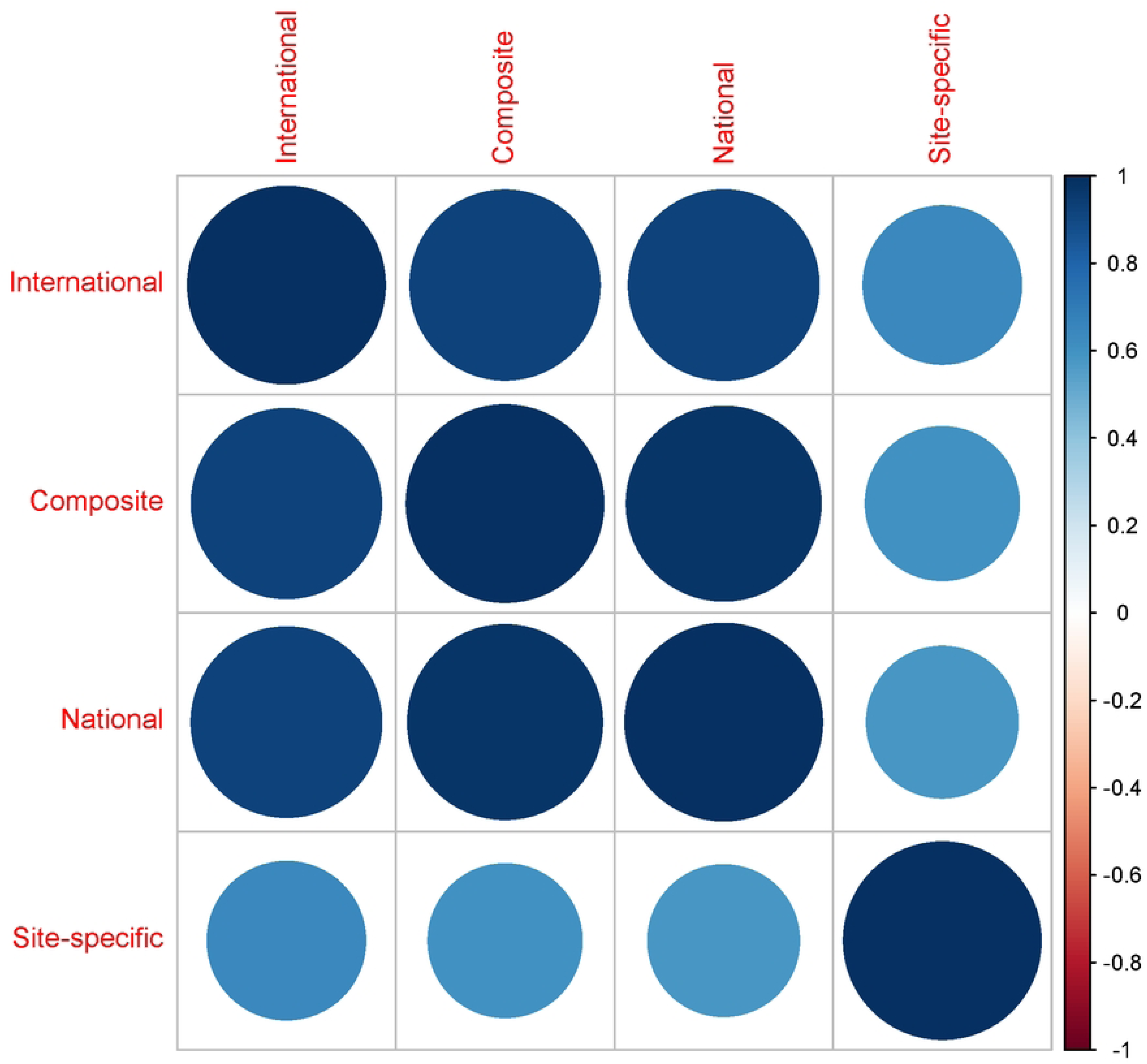
Correlation matrix between the four wealth indices. The bigger and darker the circle, the higher the correlation. Blue colour shows positive correlation

The ‘international’ and site-specific indices had a prediction accuracy of >0.55 across the three SES outcomes: high educational attainment, food security, and high household income, with the ‘international’ wealth index had a better prediction of high educational attainment: 0.67 versus 0.58. The national-level and national composite had the best prediction accuracy (0.80) of household income but with average prediction accuracy (0.50) of food security (Table 4).

**Table 4:** Accuracy of the four indices in predicting high educational attainment, food security and high household income.

| **Index** | **High educational attainment** | **Food secure** | **High household income** |
| --- | --- | --- | --- |
| **‘International’** | 0.67 | 0.56 | 0.69 |
| **National ‘composite’** | 0.59 | 0.50 | 0.80 |
| **National** | 0.58 | 0.51 | 0.80 |
| **Site-specific** | 0.58 | 0.56 | 0.70 |
| High household income was defined as the middle and high-income categories (described above). High educational attainment was defined as having secondary or tertiary education. Food security was defined as a response of 'No' to all five questions described above. |  |  |  |

The ‘international’ index was considered the best and final wealth index for project analyses for three reasons: (1) The ‘international’ index had moderate prediction accuracy (>0.55) across all three SES measures used for validation; 2) The ‘international’ index was generated in the same standardised way across the countries, allowing direct between-country comparisons; (3) The ‘international’ index had a very strong correlation with national-level and composite indices, where they were available in The Gambia and South Africa, demonstrating internal consistency.

The distribution of the ‘international’ index tertiles across the five sites had face validity; wealth index tertiles were equally distributed in urban Gambia and Zimbabwe, with more adults having a low wealth index in rural settings, (∼50%) in rural Gambia and rural South Africa, whilst a larger proportion of adults (55%) were in the middle wealth index tertile in urban South Africa (Figure 3).

**Figure 3:**
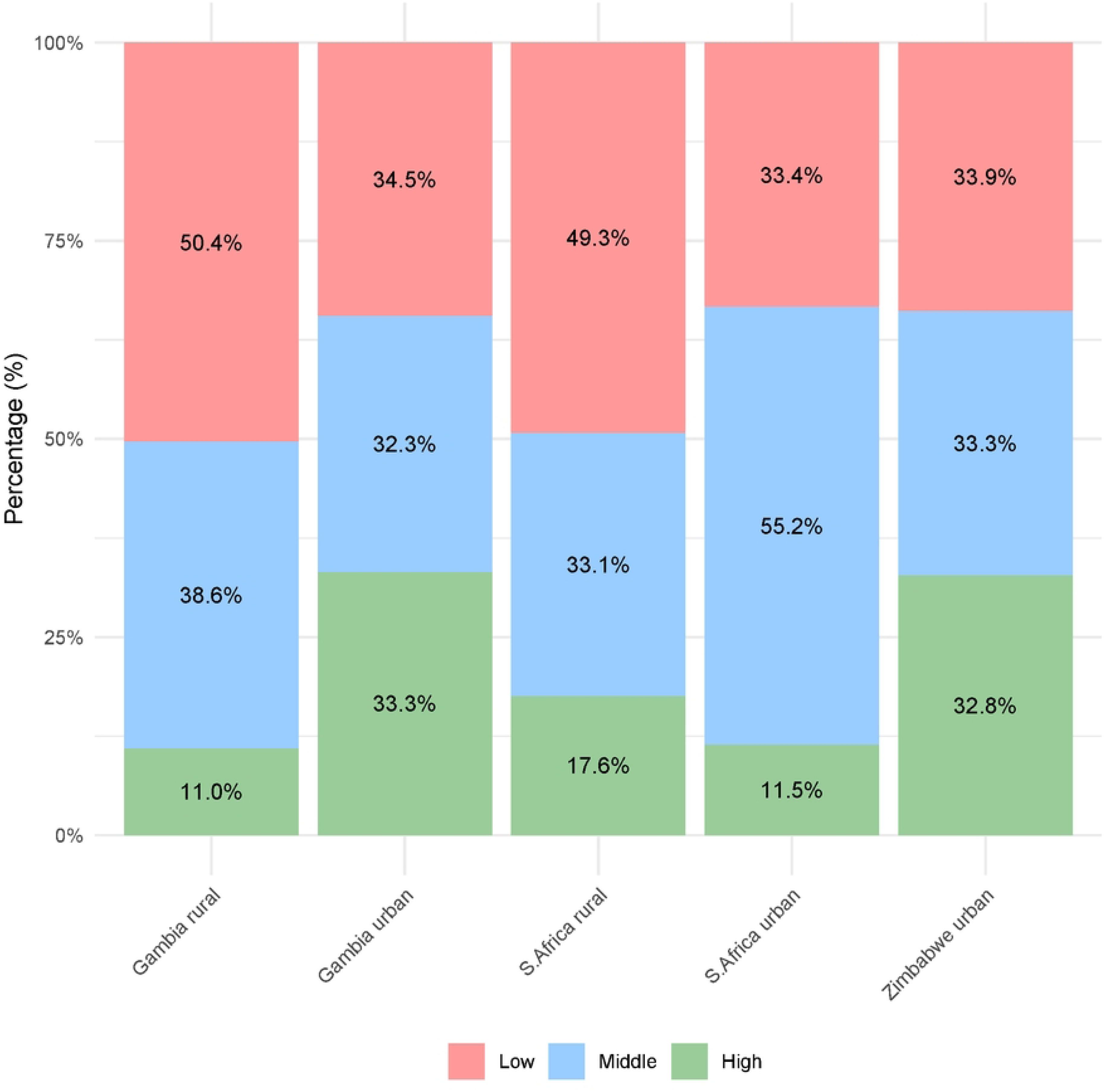
Distribution of ‘international’ wealth index tertiles across the five sites

In all five sites, as household wealth increased, quantified by the ‘international index’, ownership of a refrigerator, television, working car/truck, tap in the house, flush toilet, decoder/satellite dish, computer, and tiled floors significantly increased (Figure 4, S1 Appendix). However, country- and site-specific differences in asset ownership were seen across the wealth index tertiles. For example, while having a working car/truck was more common in urban compared to rural sites, the proportion of urban Gambian adults in the high wealth index who owned a working car/truck (50.1%) was logically lower than in urban South Africa (82.0%) (S1 Appendix).

**Figure 4:**
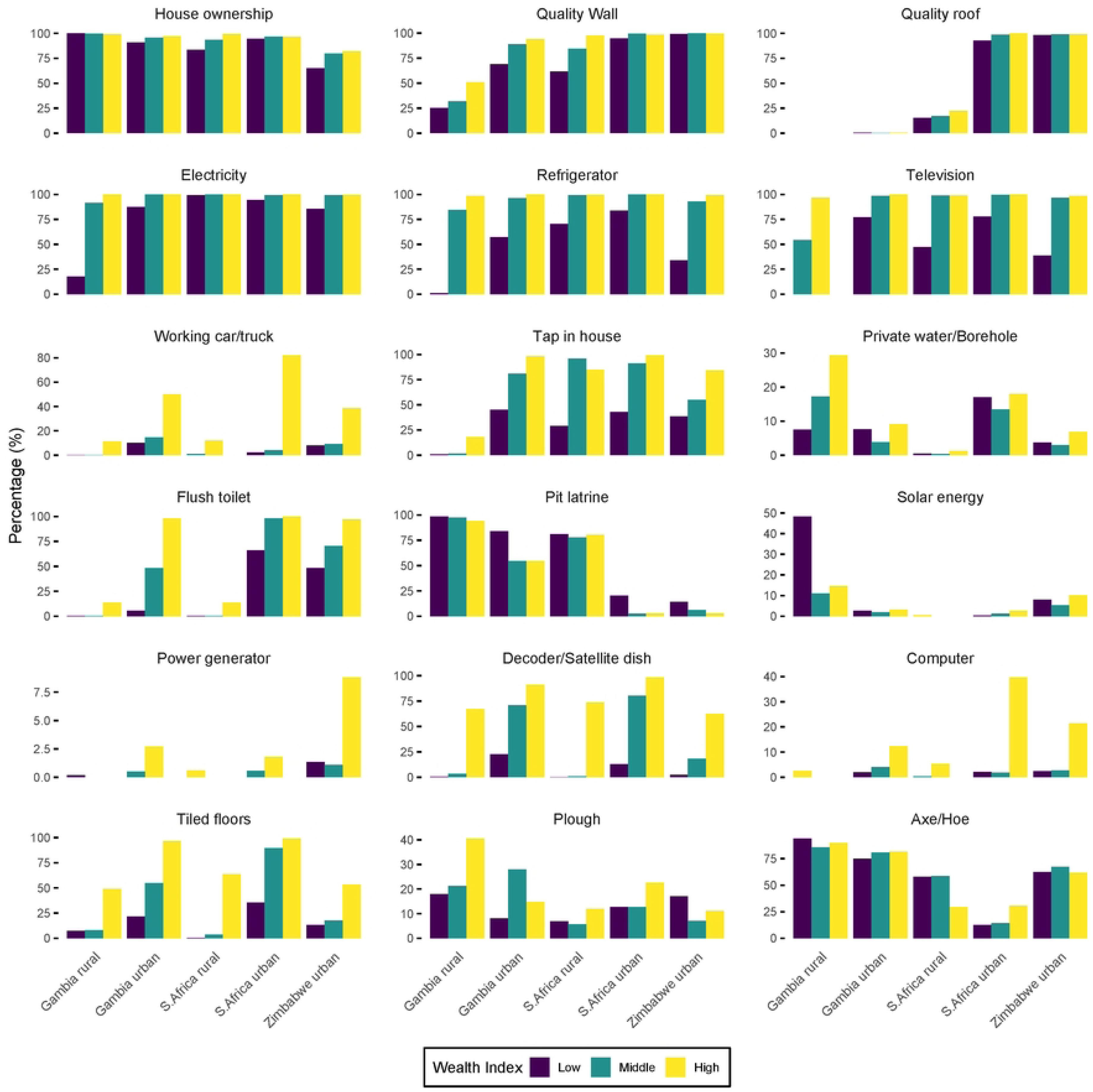
Distribution of house ownership, the quality of roof and wall materials, and 18 household assets according to the ‘international’ wealth index tertiles, across the five study sites

Finally, the ‘international’ index was validated by its associations with other SES measures: household income, educational attainment and food insecurity. Generally, greater self-reported household income was associated with an increasing wealth index in all five sites (Figure 5; S1 Appendix). Higher educational attainment was associated with a greater wealth index in all the sites, albeit the association was weak in South Africa. Except for rural Gambia, food security was associated with greater wealth in the other sites, albeit weakly in rural South Africa.

**Figure 5:**
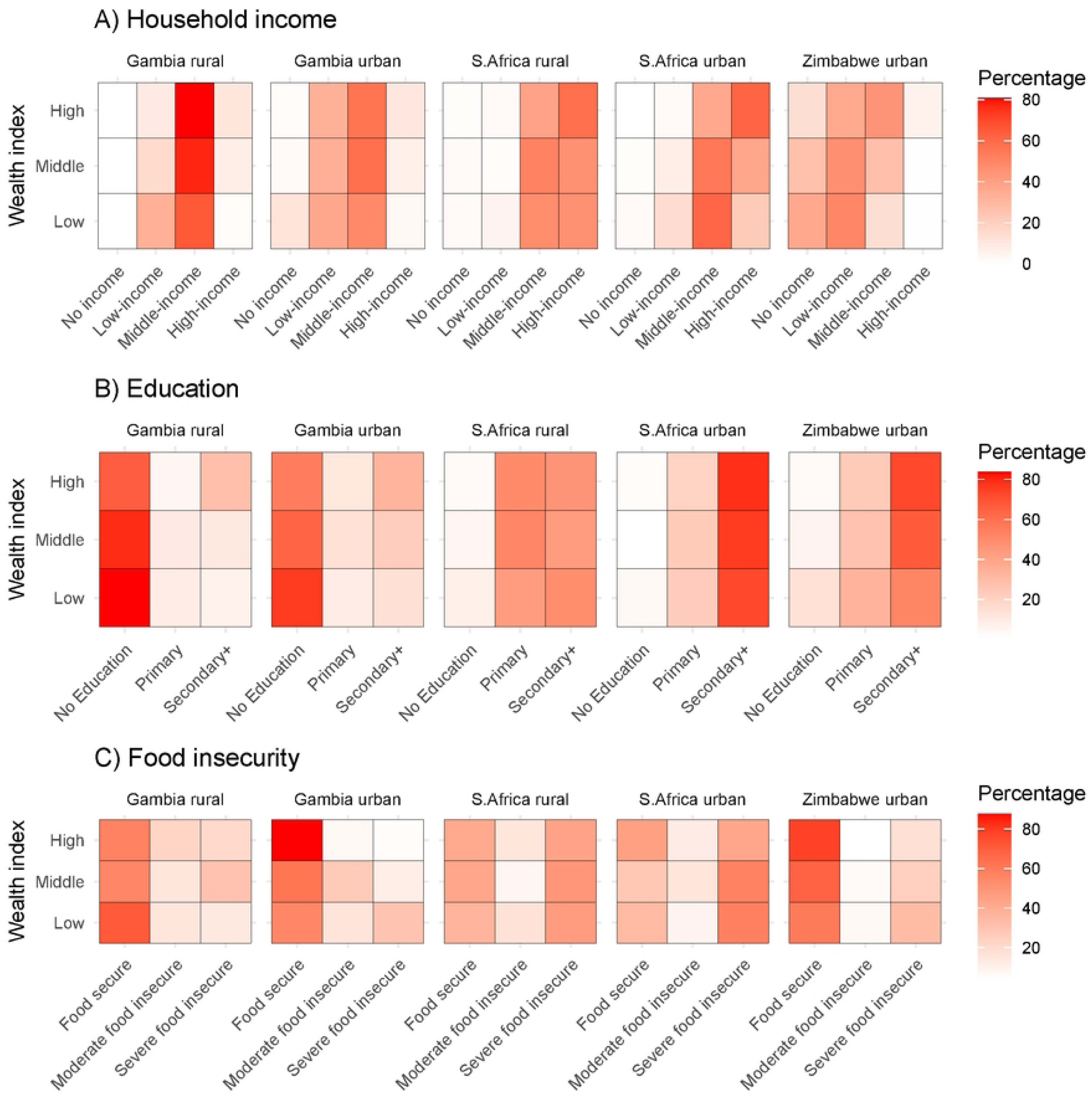
Distribution of household income, educational attainment, and food insecurity level according to ‘international’ wealth index tertiles, across the five study sites. Household income was categorised as no income, low-income (≤4000 Gambian Dalasi, ≤100 US dollars in Zimbabwe, ≤1000 South African Rand), middle-income (40,001-16,000 Gambian Dalasi, $101-500 US dollars in Zimbabwe, R1,001-3,000 South African Rand and high-income (>16,000 Gambian Dalasi, >$500 US dollars in Zimbabwe, >R3,000 South African Rand).

### Reflections on wealth measurement

Table 5 shows the wealth measurement considerations during the selection of assets, data collection, and analysis based on reflective workshops. Themes that emerged included the importance of the frequency of an asset so that it is differential (i.e., not ubiquitous and not rare); rural-urban specificity (e.g., livestock ownership may be a good measure of wealth in rural settings but not urban); and the need to remove outdated assets from the list (e.g. fuel-powered generators are largely being replaced by solar panels). Understanding the context of asset ownership was judged important in determining whether it is a measure of wealth, e.g., charitably provided pit latrines or an illegal electricity connection, which do not indicate wealth. During questionnaire design and data collection, it was felt important to consider the context of asset ownership: different types (e.g. basic telephone versus smartphone), condition (e.g., broken versus renovated), and communally versus privately owned (e.g. a borehole), which all imply different levels of wealth. Furthermore, having an asset (e.g., electricity connection) but not using it (e.g., due to unpaid bills) could imply lower wealth.

**Table 5:** Wealth measurement considerations during selection of assets, data collection and analysis.

| Consideration | Sub-themes in consideration | Explanation and examples |
| --- | --- | --- |
| **Selection of assets**<br>• *Consult the local team who understand current context when selecting assets*<br>• *Develop questionnaire incorporating feedback from consultations*<br>• *Consider similar and/or different assets for rural and urban settings*<br>• *Pilot questionnaire before data collection* | How common is an asset | • Assets selected to measure wealth should not be too rare or too common; they must have differential ability |
|  | Rural-urban differences | • While multi-site (rural and urban) studies may want to use the same questionnaire, measures of wealth may be different at sites, e.g.,<br>○ Livestock may be a good measure of wealth in rural settings<br>○ Electronics may be a good measure of wealth in urban settings |
|  | Out-dated assets | • Use of the same questionnaire over time risks including assets that have become out-of-date and are no longer indicative of wealth<br>○ In many settings, manual ploughing has been replaced with the mechanization of farming; plough ownership is no longer informative<br>○ Power generators may have been replaced by solar panels |
|  | Origin of asset | • Understand how the asset was acquired<br>○ Societal provision of an asset to an area, for community use, means that the asset is not individually differential, e.g. free government housing, neighbourhood latrines built by a non-governmental organisation<br>○ Some households may have an illegal electricity connection, indicating lower wealth |
|  | Combination of assets | • Having two assets that serve the same purpose could imply wealth<br>○ Having tap water as well as private water or a borehole<br>○ Having electricity as well as solar energy<br>• Having a flush toilet and pit latrine in an urban setting may mean a bigger house compound and more wealth |
|  | ‘Ubuntu’ practices (sharing in a community) | • If the asset is not frequently used, borrowing from neighbours can be common, and a lack of ownership is not necessarily indicative of less wealth<br>○ In urban settings, neighbours may share hoes as they are occasionally used to work on small backyard gardens |
| **Granularity in data collection**<br>• *For selected assets, consider with the local team on all possible* | Type of asset | • A simple inquiry about owning an asset overlooks asset types that indicate wealth<br>○ Phone ownership may be high, but different phone types may be differential (e.g., smartphone) |
| *types that could reflect wealth*<br>• *Consider how best and appropriate to ask for details for each asset*<br>• *Pilot the questionnaire before data collection* |  | ○ Corrugated iron roofs may be common, but some types could indicate wealth |
|  | Condition and current use of the asset | • The condition of the asset is often important<br>○ Houses may have been built at the same time and with similar material, but renovation may reflect wealth<br>○ Households may have flush toilets, but for some flushing is manual (i.e. using a bucket)<br>• Ownership without use may indicate less wealth<br>○ Houses may have an electricity connection but are not using it due to cost, or disconnection following unpaid bills |
|  | Communal or individual ownership | • Consider whether an asset is communally or privately owned<br>○ Sharing a communal borehole may indicate less wealth, while owning a private borehole indicates greater wealth |
| **Analysis**<br>• *In consultation with the local team, consider combining assets serving the same purpose as an indicator of wealth*<br>• *Use rigorous statistical approaches to construct the wealth index*<br>• *Validate the constructed wealth index against the assets collected*<br>• *Share findings and get insights from the local team* | Rigorous statistical approach | • Construct the wealth index using rigorous approaches, but also in consultation with the local team<br>• Consider integrating local insights during analysis |
|  | Validate the developed wealth index | • Validate developed wealth index against all assets used and other measures of SES such as educational attainment, income, and food security<br>• Check for face validity with the local team on trends of wealth tertiles and asset ownership observed |

During the analysis, it was considered important to take rigorous approaches to wealth index generation and validate the derived index against other SES measures (e.g., educational attainment, income). It was considered important to consult local fieldwork experts during the whole process, incorporate their insights, and use rigorous statistical approaches.

Regarding the use of self-reported household income, workshop attendees felt that it was at risk of bias from multiple sources:

- Individuals may not know the income of other household members to accurately compute a total household income
- Interviewees may expect monetary aid from participating in the research study and hence under-report their income
- Informal- and self-employment mean fluctuating incomes
- Similar incomes, but from different sources (e.g. social security or charitable grant versus employment income) may imply different levels of wealth
- Income may be viewed as only that coming from formal employment and therefore informal and/or grant income may be under-reported
- Undeclared informal income may not be self-reported, for concern over undisclosed tax liability.

Finally, workshop attendees discussed potential measures of wealth for future studies, listed in Textbox 2.

#### Textbox 2: Variables not used in the current study that could be considered in future studies for wealth measurement

**Potential measures of wealth for future studies**

***Urban and rural settings:***

- Wi-fi internet connection
- Mobile phone type, e.g. smartphone
- Employing a house helper
- Electronics (electric kettle, iron, oven, washing machine)
- House air conditioning
- Built-in cupboards and wardrobes
- Water storage tank
- Electric fences/alarm system/high durable walls

***Rural settings only:***

- Livestock ownership
- Wheelbarrow ownership

## Discussion

This study derived and compared four wealth indices by applying PCA to 18 variables describing house ownership, wall and roof material quality and household assets, based on data from more than 5000 adults living in urban and rural settings across three African countries. Reflection on data collection for wealth measurement informed recommendations for future studies. The best wealth index, out of the four generated, was significantly associated with higher ownership of eight assets, i.e., refrigerator, television, working car/truck, tap in the house, flush toilet, decoder/satellite dish, computer and tiled floors, in all five sites. Furthermore, wealth was associated with higher household income, higher educational attainment and food security, with site-specific differences in these associations.

Taken together, we recommend that when designing study questionnaires, researchers should work with local teams to understand the context in terms of; (a) frequency of asset ownership in rural and urban settings, (b) outdated and emerging indicative assets, (c) whether any charitable/state programmes provide/subsidize assets, (d) indicative asset combinations, (e) the potential for individually vs. communally owned assets, (f) types and the working condition of assets. Internal validity checks should be pre-planned, using other SES measures such as educational attainment and food insecurity; hence, these data should be collected. Finally, future studies should explore the utility of ‘emerging’ assets to measure wealth, such as Wi-Fi internet connection.

Similar to the International Wealth Index (IWI), this study found good and valid measures of wealth in all sites to be: refrigerator, television, car, tap in the house (denoting high-quality water source in IWI), flush toilet (denoting high-quality toilet facility in IWI), tiled floors (denoting high-quality floor material), decoder/satellite dish, and computer (both denoting expensive utensils)(13). Understandably, there were country and site-specific differences in ownership of these assets across the wealth index tertiles. For example, car ownership was less common in the high wealth tertile group in urban Gambia compared to the same category in urban South Africa. Such differences are likely partly explained by wealth differences between the two countries, with World Bank estimates showing that South Africa has a substantially higher Gross Domestic Product (GDP) per capita compared to The Gambia: 6,253.2 versus 843.8 US dollars(20). Previous literature has reported that differences in asset ownership are influenced by several factors, including rural-urban setting, the local association of wealth to certain assets, the utility of assets, and community infrastructure (e.g. electricity and water supplies) (4, 6, 21). The recommendation made on the need to consult local fieldwork teams when selecting the most wealth-relevant assets for a study population has been echoed and implemented by other researchers, including through the use of interviews, focus group discussions and observations to generate an asset list for wealth measurement(4). Such consultation can identify the most relevant assets, improving wealth estimation and reducing data collection burden, by selecting a small selection of differential assets.

Apart from careful selection of assets, the level of detail needed for the asset during data collection must also be specified to measure wealth more accurately. This includes the type of asset, condition of asset, and private or communal ownership. Importation of less expensive assets to Africa, mainly from China and Asia(6, 22), means that assets could be common but with lower values. Therefore, we recommend that, where relevant, data are collected on different types of the same asset, and when relevant, data on the condition (i.e. is it working?). Implementing these recommendations could generate categorical rather than binary variables, which may increase data collection time; however, focusing just on relevant assets would offset time delays.

Finally, after careful data collection, rigorous statistical approaches should be used to generate the wealth index. We used the PCA approach, which has been commonly used to generate wealth indices in low- and middle-income countries(23). Nevertheless, limitations in this approach have been documented (e.g., the principal component may explain only a low proportion of variance in asset data), and alternative approaches and statistical considerations (e.g., multiple correspondence analysis) have been discussed in the literature (3, 4, 6, 9).

Regardless of the approach used, validating the derived wealth index against other SES measures is important to ascertain internal validity. In our analysis, we found household income, educational attainment and food security were associated with increasing wealth in line with other studies(6, 12, 24). However, it is important to note that relationships between these SES measures are complex; each measure is distinct and it may be inappropriate to use these interchangeably(24, 25), such that the wealth index may not always be a more valid estimate of wealth than other measures such as income(21). For example, in the current study, none of the four indices achieved an 80% prediction accuracy for the other SES measures (high educational attainment, food security and high household income).

The strengths of this paper are the use of a large sample spanning rural and urban sites in three diverse African countries, the generation of four wealth indices, their comparison and validation of a final wealth index. Furthermore, quantitative analyses were supplemented by qualitative reflective workshop analyses. Using nine assets to generate the ‘international’ wealth index is pragmatic and practical and could minimise the data collection burden.

However, data on a greater number of assets, particularly rural-specific assets, was a potential limitation of this analysis. Furthermore, although PCA is a commonly used approach, we did not appraise the generated wealth index against other statistical approaches. Nevertheless, the generated wealth index showed an expected relationship with other SES measures such as household income. The methodology used in this study can be used, by multi-country studies, to inform the generation of wealth indices across geographical sites and countries when standardised and comparable indices cannot be generated, such as the IWI.

## Conclusion

Ownership of a refrigerator, television, working car/truck, tap in the house, flush toilet, decoder/satellite dish, computer and tiled floors were associated with greater wealth in all five sites albeit with site and country differences. Accuracy of wealth measurement for multi-country and multi-site studies using asset-based indices needs unambiguous, differential, and clearly defined asset lists. Assets selected should be country- and site-relevant and research teams should consider asset frequency, dropping outdated assets, and the potential inclusion of emerging indicative assets such as Wi-Fi internet connection, whether assets can be accessed for free/ by subsidy, or whether individually or communally owned, and provide sufficient detail on asset type and working condition. Rigorous statistical approaches to generate indices should be used, including internal validation of the generated indices. Throughout the process, insights from local fieldwork experts should be sought to inform selection, data collection, and analysis.

## Data Availability

Data are available upon reasonable request. Researchers can access participant-level data via data.bris, provided ethical approvals are in place.

## Ethical statement

Ethical approval was obtained in all the study countries. The Gambia: The Gambia Government/MRC Unit The Gambia, LSHTM Scientific Coordinating Committee and Ethics committee (22/04/2021 ref 22975); Ministry of Health (20/08/2021 ref DDHS/AD/2021/08(MTN27)). Zimbabwe: The Medical Research Council of Zimbabwe (14/07/2021 ref MRCZ/A/2706); The Biomedical Research and Training Institute (19/02/2021 ref AP161/2021); Sally Mugabe Central Hospital (29/01/2021 ref HCHEC/ 250121/06); The University of Zimbabwe College of Health Sciences and the Parirenyatwa group of hospitals (25/02/2021); Harare City Health (27/01/2021); The Research Council of Zimbabwe (RCZ, 14/07/2021 refs 04246 and 04248). South Africa: The University of KwaZulu-Natal’s Biomedical Research Ethics Committee (BREC, WP1 21/08/2021 BREC/00002513/2021), WP3 10/03/2021 BREC/00002125/2021, WP4 21//08/2021 BREC/00002423/2021) and the University of Witwatersrand’s Health Research Ethics Committee (Medical, WP5 20/08/2021 ref R14/49).

All participants provided written informed consent before data collection.

## Acknowledgements

The authors would like to acknowledge all participants who took part in the study, and the fieldworkers, clinical and support staff who facilitated data collection.

## Supplementary material caption

## S1 Appendix

